# An empirical investigation into the impact of winner’s curse on estimates from Mendelian randomization

**DOI:** 10.1101/2022.08.05.22278470

**Authors:** Tao Jiang, Dipender Gill, Adam S. Butterworth, Stephen Burgess

**Affiliations:** BHF Cardiovascular Epidemiology Unit, Department of Public Health and Primary Care, University of Cambridge, Cambridge CB1 8RN, UK; Department of Epidemiology and Biostatistics, School of Public Health, Imperial College London, London, UK; Chief Scientific Advisor Office, Research and Early Development, Novo Nordisk, Copenhagen, Denmark; British Heart Foundation Centre of Research Excellence, University of Cambridge, Cambridge CB2 0QQ, UK; National Institute for Health Research Blood and Transplant Research Unit in Donor Health and Genomics, University of Cambridge, Cambridge CB1 8RN, UK; Health Data Research UK Cambridge, Wellcome Genome Campus and University of Cambridge, Cambridge CB10 1SA, UK; MRC Biostatistics Unit, University of Cambridge, Cambridge CB2 0SR, UK

## Abstract

**Introduction:** Genetic associations for variants identified through genome-wide association studies (GWAS) tend to be overestimated in the original discovery dataset; as if the association was underestimated, the variant may not have been detected. This bias, known as winner’s curse, can affect Mendelian randomization estimates, but its severity and potential impact is unclear.

**Methods:** We performed an empirical investigation to assess the potential bias from winner’s curse in practice. We considered Mendelian randomization estimates for the effect of body mass index (BMI) on coronary artery disease risk. We randomly divided a UK Biobank dataset 100 times into three equal-sized subsets. The first subset was treated as the “discovery GWAS”. We compared genetic associations estimated in the discovery GWAS to those estimated in the other subsets for each of the 100 iterations.

**Results:** For variants associated with BMI at *p*<5×10^−8^ in at least one iteration, genetic associations with BMI were up to five-fold greater in iterations where the variant was statistically significantly associated with BMI compared to its mean association across all iterations. If the minimum p-value for association with BMI was *p*=10^−13^ or lower, then this inflation was less than 25%. Mendelian randomization estimates were affected by winner’s curse bias. However, bias did not materially affect results; all analyses indicated a deleterious effect of BMI on CAD risk.

**Conclusions:** Winner’s curse can bias Mendelian randomization estimates, although its practical impact may not be substantial. If avoiding sample overlap is infeasible, analysts should consider performing a sensitivity analysis based on variants strongly associated with the exposure.

## Introduction

Mendelian randomization is an epidemiological approach in which genetic variants are used as instrumental variables to assess the existence and potential magnitude of the causal effect of an exposure on an outcome (1, 2). Due to the nature of Mendelian inheritance, there is inherent randomness in the transmission of genetic variants from parent to offspring (3, 4). This randomness has been shown to hold approximately at a population level for many common genetic variants (5, 6). It is therefore possible to treat genetic variants associated with a modifiable trait as unconfounded proxies for the effect of altering that trait, thus mimicking treatment allocation in a randomized controlled trial (7). This makes Mendelian randomization a flexible and credible approach to make causal inferences for a wide range of exposure—outcome pairs. With the rapid expansion of publicly available summary statistics from genome-wide association studies (GWAS) over the recent decade (8), Mendelian randomization analyses using such statistics have become increasingly popular (9).

The concept of winner’s curse originates from auctions where multiple bidders each have different private estimates on the value of the item for sale (10). Under the assumption that all bids are unbiased estimates of the true value of the item but with error, the winner’s bid price will generally be an overestimate of the true value, as the final selling price is the highest bid. This upwards bias in the winner’s bid compared to the true value is known as winner’s curse. In GWAS, the estimated associations of the reported significant variants for a trait are likely to be upwards biased, as they are identified based on a statistical significance threshold – only the ”winning” variants are reported as significant (11). While winner’s curse bias will be most severe for associations with the trait under investigation, genetic associations with any variable correlated with the trait will also tend to be over-estimated. In particular, in the context of Mendelian randomization, genetic associations with an outcome will likely also be over-estimated in the discovery GWAS dataset for an exposure, as exposures and outcomes are typically associated due to confounding.

This bias can have a direct impact on Mendelian randomization estimates calculated using association estimates derived from the discovery GWAS. With a single genetic variant, the Mendelian randomization estimate can be expressed as the ratio of the genetic association with the outcome divided by the genetic association with the exposure (12). With multiple genetic variants, the standard combined estimate (the inverse-variance weighted estimate) is a weighted mean of these ratio estimates calculated for each variant (13). Hence, winner’s curse in the *exposure* association estimates would be expected to result in a *deflation* in the Mendelian randomization estimate, whereas winner’s curse in the *outcome* association estimates would be expected to result in an *inflation* in the Mendelian randomization estimate.

Winner’s curse can be alleviated by selecting genetic variants and estimating genetic associations in non-overlapping datasets. However, it may not be possible to find distinct sets of summary statistics from independent datasets for the same trait. The degree of bias from winner’s curse depends on various factors, and is typically worse for genetic variants with associations close to the statistical significance threshold. Several methods have been proposed to correct for winner’s curse bias (14–18). However, these methods can be overly conservative and result in a loss in power, or assume an underlying distribution that may not hold.

A related phenomenon in Mendelian randomization is weak instrument bias, which arises due to chance associations of the genetic variants with confounders (19, 20). Even if a genetic variant is a valid instrumental variable (i.e. has no true association with confounders), correlations of the variant with confounders will not be exactly zero. If a genetic variant is strongly associated with the exposure, then bias due to chance associations with the confounders is negligible. However, if a genetic variant is weakly associated with the exposure, then bias due to chance imbalances in the distribution of a confounder can be non-negligible. For a *one-sample* Mendelian randomization analysis, in which the same dataset is used for estimating the genetic associations with the exposure and the outcome, these chance correlations affect associations with both the exposure and the outcome in a related way, and bias (known as weak instrument bias) is *towards the observational association* between the exposure and outcome. For a *two-sample* Mendelian randomization analysis, in which genetic associations with the exposure and outcome are obtained from independent samples, these chance correlations will differ between the datasets and so affect associations with the exposure and the outcome independently, and weak instrument bias is *towards the null* (21). The magnitude of weak instrument bias depends on the instrument strength, which can be estimated as the F statistic from regression of the exposure on the genetic variants (22, 23).

Both winner’s curse and weak instrument bias are finite-sample biases, in that they reduce towards zero when the sample size gets large. However, while the expected magnitude of bias due to weak instruments can be approximated (and is typically slight when the genetic variants are associated with the exposure at a genome-wide level of statistical significance), the expected magnitude of bias due to winner’s curse in a Mendelian randomization investigation is typically unclear. This is important to study, as any analysis must balance concerns about winner’s curse and weak instrument bias against other biases, such as bias from instrument invalidity due to pleiotropy.

Here, we provide an empirical investigation into the magnitude of bias from winner’s curse and weak instruments in an applied Mendelian randomization analysis considering the effect of body mass index (BMI) on coronary artery disease (CAD) risk. We picked this example because there is previous evidence supporting a causal relationship (24, 25), and several hundred genetic variants have been found to be associated with BMI in previous GWAS, meaning that we can compare genetic association estimates for a large number of variants. We took data from UK Biobank, which we randomly split into three equal-sized subsets to consider different scenarios in which the genetic variants were chosen based on their associations in one subset (the “discovery GWAS”), and associations with the exposure and outcome were either estimated in the same subset or a different subset. We repeated this splitting procedure to investigate the distribution of Mendelian randomization estimates. To compare the magnitude of bias in different situations, we also considered Mendelian randomization estimates obtained only using genetic variants that were statistically strongly associated with the exposure, as well as estimates obtained only using genetic variants that were not fully consistent in their association with the exposure across datasets. We conclude by discussing the relevance of these findings for bias in other Mendelian randomization investigations, and the implications for how Mendelian randomization investigations should be performed.

## Methods

### STUDY POPULATION AND OUTCOMES

All analyses were performed in the UK Biobank dataset, a prospective cohort study of around 500,000 UK residents aged 40 to 69 years. We took data on 367,644 unrelated participants of European ancestries, as ascertained by a mixture of self-report and genetic information following quality control procedures previously described (26). BMI was calculated as weight in kilogrammes divided by height in metres squared. CAD was defined using International Classification of Diseases, Tenth Revision (ICD-10) codes as ICD-10 code I21-I25 or self-reported data from interview with a nurse practitioner.

### DISCOVERY GWAS AND OBTAINING MENDELIAN RANDOMIZATION ESTIMATES

We split UK Biobank participants into three equal-sized groups at random, which we refer to as Group A (the discovery GWAS group), Group B, and Group C. Using data from Group A, we performed a genome-wide association analysis for BMI adjusting for age, sex and the first ten genomic principal components. Variants were filtered for a minor allele frequency of >0.01% and an INFO score (an imputation quality metric) of >0.4 (27). Variants passing these filters and reaching the conventional genome-wide significance threshold of *p* < 5×10^−8^ were selected. In order to ensure that selected variants are mutually independent, we clumped the selected variants into loci using a distance threshold of 1 megabase and a correlation threshold of r^2^<0.01 estimated within Group A, to ensure no two selected variants are too close or too highly correlated with each other. We selected the variant with the smallest p-value in each locus to create our discovery set.

For each variant in our discovery set, we estimated its genetic association with BMI in each of Groups A, B, and C by linear regression adjusting for age, sex and the first ten genomic principal components. We also calculated genetic associations with CAD risk in each group by logistic regression with the same covariate adjustment. We combined these summarized genetic association estimates across variants to obtain Mendelian randomization estimates using the random effects inverse-variance weighted method (28). This estimate represents the log odds ratio for CAD per 1 kg/m^2^ higher genetically-predicted BMI. This entire process was repeated for 100 iterations taking different random splits of the original dataset in each iteration.

### ANALYSIS SET-UP

We considered five scenarios, corresponding to different situations in which winner’s curse and weak instrument bias should or should not occur (**Table 1**). We use the term “overlap” to indicate that genetic associations are estimated in the same participants as the discovery GWAS (Group A), and one-sample or two-sample to indicate whether genetic associations with the exposure and outcome are estimated in the same or different groups respectively.

**Table 1:** Summary of scenarios considered in the empirical analysis. In each scenario, Group A was used as the discovery GWAS.

| Scenario | Group genetic associations<br>estimated in for... |  | Winner's curse affects genetic<br>association estimates for... |  | One-sample? |
| --- | --- | --- | --- | --- | --- |
|  | Exposure | Outcome | Exposure | Outcome |  |
| 1 | B | C |  |  |  |
| 2 | A | B | ✓ |  |  |
| 3 | B | A |  | ✓ |  |
| 4 | B | B |  |  | ✓ |
| 5 | A | A | ✓ | ✓ | ✓ |

1. **No overlap, two-sample**: genetic associations with BMI are estimated in Group B and genetic associations with CAD in Group C, i.e. we have three distinct datasets for discovery, exposure associations, and outcome associations. This represents a scenario in which winner’s curse will not occur, and weak instrument bias will be towards the null. We treat this as the reference scenario for comparison purposes.
2. **Overlap with exposure, two-sample**: genetic associations with BMI are estimated in Group A and genetic associations with CAD in Group B, i.e. we have overlapping discovery and exposure datasets. Winner’s curse will affect genetic associations with BMI, but not with CAD risk, so Mendelian randomization estimates should be deflated. Weak instrument bias will be towards the null.
3. **Overlap with outcome, two-sample**: genetic associations with BMI are estimated in Group B and genetic associations with CAD in Group A, i.e. we have overlapping discovery and outcome datasets. Winner’s curse will affect genetic associations with CAD risk, but not with BMI, so Mendelian randomization estimates should be inflated. Weak instrument bias will be towards the null.
4. **No overlap, one-sample**: genetic associations with BMI and CAD were both estimated in Group B, i.e. we have no overlap between the discovery and estimation datasets, but genetic associations with the exposure and outcome were obtained in the same dataset. Winner’s curse will not occur, and weak instrument bias will be towards the observational association between BMI and CAD risk (which is positive).
5. **Overlap with exposure and outcome, one-sample**: genetic associations with BMI and CAD were both estimated in Group A, i.e. all analyses were performed on the same dataset. This represents a scenario in which winner’s curse will occur for genetic associations with both BMI and CAD risk, so Mendelian randomization estimates will be subject to both inflation and deflation; it is not clear what the net effect will be. Weak instrument bias will be towards the observational association between BMI and CAD risk.

In addition to assessing winner’s curse in the Mendelian randomization estimates, we also considered the amount of winner’s curse in the genetic associations for individual variants, defined as the percentage difference in the mean beta-coefficient across iterations for which the association was significantly associated with BMI (at *p* < 5×10^−8^) divided by the mean beta-coefficient across all 100 iterations:

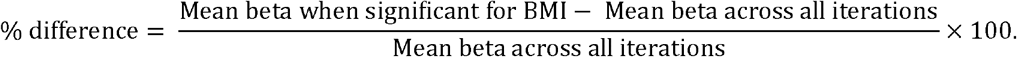

We define the absolute difference similarly:

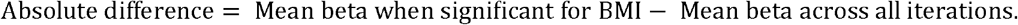

Note that these measures are defined for genetic associations with BMI and with CAD risk, but in both cases, statistical significance is judged based on the genetic associations with BMI. This reflects that in a Mendelian randomization investigation, variants are selected based on their associations with the exposure, not the outcome.

In secondary analyses, we considered two further strategies for selecting variants. First, we considered a stricter statistical significance threshold for variant selection of *p* < 5×10^−11^. Only variants meeting this threshold were included in the Mendelian randomization analyses. Second, we noted which genetic variants were selected into the discovery set for all of the 100 random splits of the original dataset. Loci from which a variant was selected in all 100 random splits were removed from these analyses. The motivation of this “no full replication” strategy is to assess the impact of winner’s curse for a trait when only variants with relatively weaker evidence of association are available.

Genetic association analyses were performed using SNPTEST v2 (29) and Mendelian randomization analyses were performed using the MendelianRandomization package (30) in R v3.6.1 (31).

## Results

The mean age of the 367,644 participants at baseline was 57.2 years, and 54.1% of participants were female. Mean BMI was 27.4 kg/m^2^, and there were 29 330 CAD events. Overall, variants in 359 loci were associated with BMI at a genome-wide significance threshold (*p* < 5×10^−8^) in Group A for at least one iteration of the random splitting procedure. Of these, 7 loci contained a variant significantly associated with BMI in all 100 iterations, whereas variants in the remaining 352 loci were significantly associated with BMI in some iterations but not others. The median number of variants selected in each iteration was 39.

Inflation of genetic associations due to winner’s curse for individual variants is illustrated in **Figure 1**. For genetic variants that had a minimum p-value for association with the exposure across iterations of around 5×10^−8^, the percentage difference for the association with BMI in iterations when they were statistically significant in their association with the exposure compared with its mean value across all iterations varied from 50% to 400%, suggesting that beta-coefficients for associations with the exposure were on average between 1.5 and 5 times too large in these datasets. For genetic variants that had a minimum p-value of around 1×10^−13^, the percentage difference varied from 10% to 25% (i.e. 1.1 to 1.25-fold inflation due to winner’s curse). A similar pattern was observed in the absolute difference in associations, which reduced in magnitude considerably as the minimum p-value decreased. Inflation was also observed in the genetic associations with CAD risk. On the percentage difference scale, several variants had inflated associations with CAD risk up to and beyond 400% (**Supplementary Figure S1**). However, some of these percentage differences were large because the mean association with CAD risk across was all iterations was close to zero. On the absolute difference scale, associations with CAD risk were less inflated than those with BMI.

**Figure 1:**
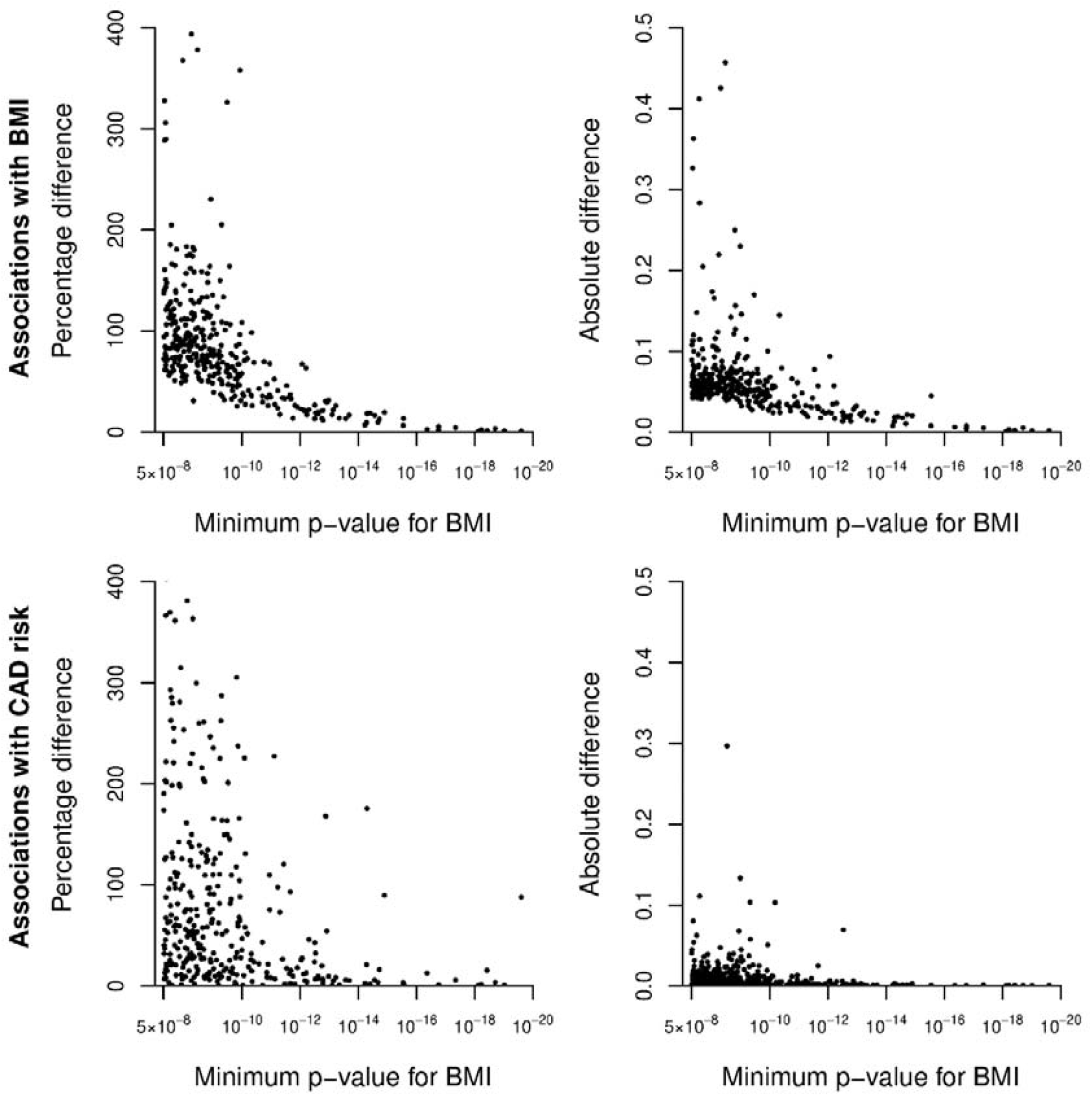
Scatter plot showing variants selected as associated with BMI in at least one iteration: the percentage and absolute differences in the mean beta-coefficient estimate in Group A between the mean value across all iterations and the mean across only those iterations for which it was significant for BMI calculated for associations with BMI (top row) and CAD risk (bottom row), plotted against its minimum p-value for BMI across iterations. Only one variant per locus is plotted. 9 variants had a minimum p-value below 10^−20^; percentage and absolute differences were close to zero for these variants. Percentage differences for associations with CAD risk exceeded 400 for 31 variants (maximum value was 6452); these points are not shown on these axes (see **Supplementary Figure S1**).

Mendelian randomization estimates are summarized in **Table 2**. In the primary analyses using all variants, all Mendelian randomization estimates were positive and had *p* < 0.05 in each iteration. The median estimate from Scenario 1 (no overlap, two-sample) was 0.0859, corresponding to an odds ratio of 1.090 per 1 kg/m^2^ increase in genetically-predicted BMI, similar to what has been observed previously (25). As expected, the median estimate from Scenario 2 (overlap with exposure, two-sample) deflated to 0.0720, and the median estimate from Scenario 3 (overlap with outcome, two-sample) inflated to 0.0950. Judging by the differences in median estimates, winner’s curse in genetic associations with the outcome affected Mendelian randomization estimates less strongly than winner’s curse in genetic associations with the exposure, although the difference was slight.

**Table 2:** Summary of primary analysis results. Estimates represent log odds ratios for coronary artery disease per 1 kg/m^2^ increase in genetically-predicted body mass index. For each scenario, we report the median and mean estimates across 100 iterations, the mean standard error of estimates, and the standard deviation of estimates. All estimates are obtained from the random-effects inverse-variance weighted method.

|  | Scenario | Median estimate | Mean estimate | Mean standard error | Standard deviation of estimates |
| --- | --- | --- | --- | --- | --- |
| 1 | No overlap, two-sample | 0.0859 | 0.0846 | 0.0158 | 0.0123 |
| 2 | Overlap with exposure, two-sample | 0.0720 | 0.0724 | 0.0164 | 0.0132 |
| 3 | Overlap with outcome, two-sample | 0.0950 | 0.0948 | 0.0204 | 0.0152 |
| 4 | No overlap, one-sample | 0.0913 | 0.0893 | 0.0204 | 0.0157 |
| 5 | Overlap with exposure and outcome, one-sample | 0.0829 | 0.0829 | 0.0201 | 0.0142 |

Comparing Scenario 1 (no overlap, two-sample) and Scenario 4 (no overlap, one-sample) shows the impact of weak instrument bias separate from winner’s curse. The median estimate from Scenario 4 was 0.0913, suggesting that the impact of weak instrument bias is less than the impact of winner’s curse in this example. Scenario 5 (overlap with exposure and outcome, one-sample) is the most complex scenario to predict, as here we have winner’s curse in the genetic associations with both exposure and outcome, plus weak instrument bias in the direction of the observational association. Winner’s curse with the exposure would be expected to deflate estimates, whereas winner’s curse with the outcome and weak instrument bias would be expected to inflate estimates. The median estimate from Scenario 5 was 0.0829, indicating that the competing biases approximately cancelled out.

Results from the secondary analyses considering different strategies for selecting variants are presented in **Figure 2**. First, we see that median estimates in Scenario 1 (no overlap, two-sample) differ slightly between the three strategies. There is no systematic reason why these differences occur, other than that they are based on different variants that may affect BMI in different ways. In this example, estimates using the strict strategy (blue boxes) are generally slightly smaller in magnitude than those from the primary strategy using all variants (red boxes), whereas estimates using the “no full replication” strategy (green boxes) are slightly larger.

**Figure 2:**
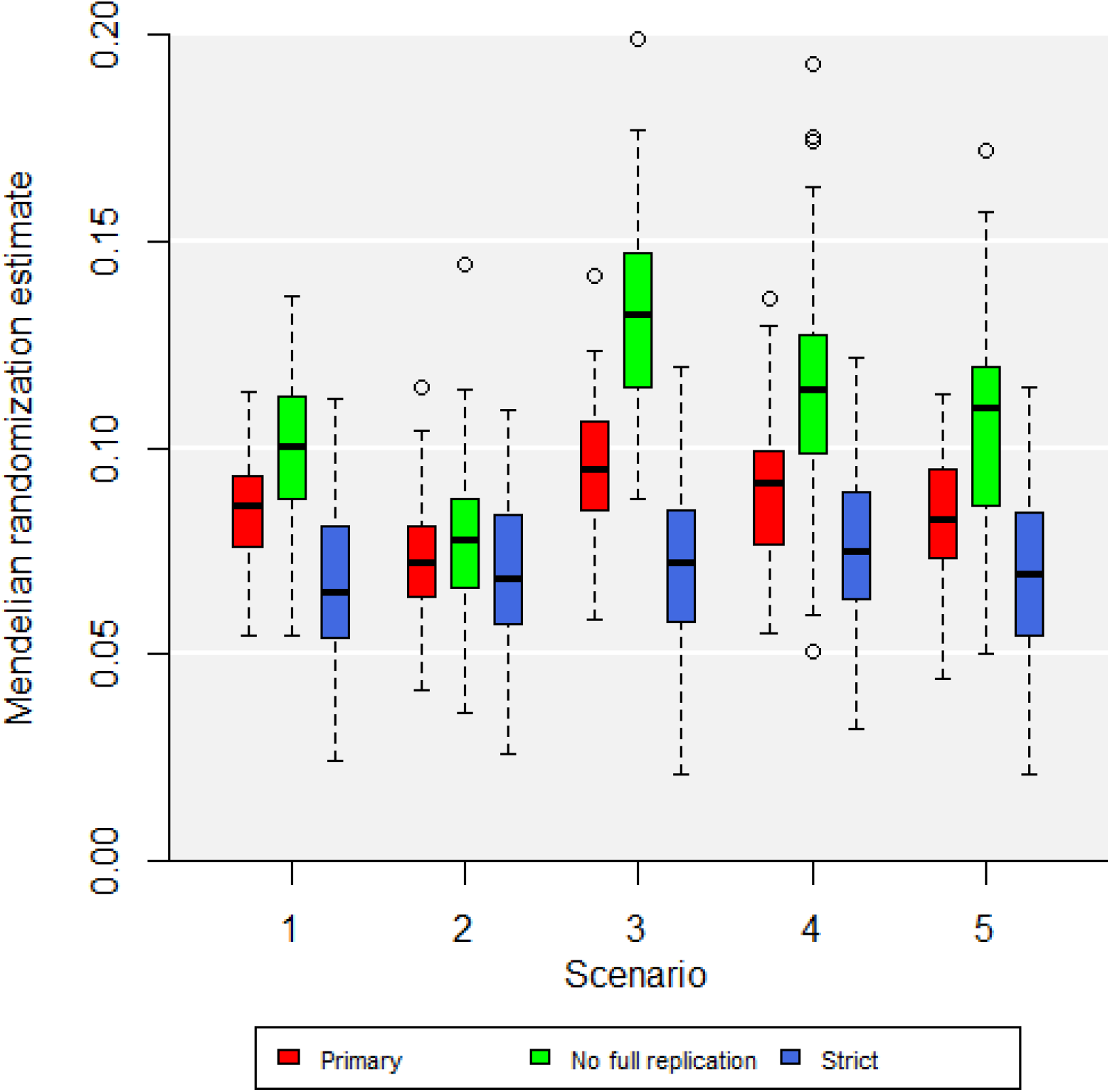
Boxplots of primary and secondary results. Estimates represent log odds ratios for coronary artery disease per 1 kg/m^2^ increase in genetically-predicted body mass index. Boxplot indicates the lower quartile, median, and upper quartile of estimates; error bars represent the range of estimates up to 1.5 times the interquartile range. Outliers outside this range are plotted separately. All estimates are obtained from the random-effects inverse-variance weighted method.

Compared with Scenario 1, the pattern of median estimates in Scenarios 2 to 4 is broadly the same for each variant selection strategy. Median estimates are deflated in Scenario 2, and inflated in Scenarios 3 and 4. The degree of variation due to bias was around 2 to 3 times larger for the “no full replication” strategy and much less for the strict strategy. In the primary and “no full replication” strategies, inflation was greater in Scenario 3 (due to winner’s curse) than in Scenario 4 (due to weak instrument bias). In contrast, in the strict strategy, inflation was slightly greater in Scenario 4 than Scenario 3, although the difference was minimal. Additionally in the strict strategy, no deflation was observed in Scenario 2. While there was some difference of median estimates in Scenario 5, this was not substantial for any of the strategies.

## Discussion

In this paper, we have explored the impact of winner’s curse bias on Mendelian randomization estimates for the effect of BMI on CAD risk. By dividing the UK Biobank study into three groups at random a large number of times, we were able to compare the distribution of Mendelian randomization estimates in scenarios where the discovery and estimation datasets were distinct, and scenarios where they overlapped. When the discovery dataset overlapped with the dataset used for estimating genetic associations with the exposure, Mendelian randomization estimates were typically deflated; whereas when the discovery dataset overlapped with the dataset for estimating genetic associations with the outcome, Mendelian randomization estimates were typically inflated.

We were able to compare the magnitude of bias from winner’s curse to bias from weak instrument bias. In the primary analysis including all genome-wide significant variants, bias from winner’s curse was greater in magnitude. Bias from both weak instruments and winner’s curse was substantially lower when only including variants associated with BMI at a stricter significance threshold, and bias was substantially greater when excluding variants consistently strongly associated with BMI. In the latter case, winner’s curse bias was again greater in magnitude than weak instrument bias. In the former case, both biases were slight, although weak instrument bias was slightly larger than winner’s curse bias. Additionally, we showed that winner’s curse for individual variant associations can be very substantial in practice, with inflation in beta-coefficients for the exposure of 50% to 400% for variants that only just achieved the GWAS significance threshold (*p* ≈ 5×10^−8^). However, the degree of bias dropped off sharply for variants associated with the exposure at a stricter significance threshold.

Despite bias from winner’s curse and weak instruments, Mendelian randomization evidence for a positive effect of BMI on CAD risk was obtained in all primary analysis iterations. While it would be unwise to conclude that winner’s curse bias is never substantial in Mendelian randomization analyses based on a single empirical analysis, some important lessons can be learned from this work. First, the magnitude of bias from winner’s curse depends sharply on the statistical strength of genetic associations. If genetic variants are strongly associated with the exposure (in terms of statistical strength of evidence), then there is little uncertainty in whether they will be selected or not from the discovery GWAS, and so winner’s curse bias is minimal. In contrast, if genetic associations with the exposure are close to the GWAS significance threshold, then bias will be more substantial. Second, bias from winner’s curse was generally greater in magnitude than weak instrument bias, but the two biases had a roughly similar order of magnitude in our empirical example.

While winner’s curse bias in genetic associations with the exposure is unwelcome, in isolation it will not lead to inflated Type 1 error rates for Mendelian randomization estimates. Although the magnitude of a Mendelian randomization estimate depends on associations with the exposure and outcome, its significance only depends on the genetic associations with the outcome. This is because the null hypothesis that the Mendelian randomization estimate is zero is achieved exactly when the genetic association with the outcome is zero; if the genetic association with the outcome is zero, then the Mendelian randomization estimate will be zero regardless of the genetic association with the exposure. Additionally, winner’s curse in outcome association estimates typically inflates Mendelian randomization estimates, which is more worrying as it can lead to false positive findings, whereas deflation in estimates is typically conservative. Hence, winner’s curse bias is a more serious problem in practice when it affects genetic associations with the outcome. Fortunately, as shown in our example, the absolute bias in genetic association estimates due to winner’s curse is less for associations with the outcome compared to associations with the exposure, as genetic variants are selected based on their associations with the exposure. However, the proportional bias can still be large.

For simplicity, we have only considered scenarios where there is either complete or no overlap between the discovery and estimation datasets. If there is partial overlap between datasets, winner’s curse bias will be less substantial. Given their international collaborative nature, many GWAS consortia have substantial overlap. Additionally, the same participants may have been recruited into multiple studies. Hence, even if genetic associations appear to come from non-overlapping datasets, participant overlap may be non-zero in practice.

The ideal situation is that Mendelian randomization analyses operate using a “three-sample” design, in which discovery GWAS, exposure associations, and outcome associations are assessed in non-overlapping datasets (32). However, in practice it is rarely feasible to find three distinct large samples of participants that are sufficiently similar to combine into a single analysis (for example, same ancestry group). Even if it is possible, the loss in sample size (and hence power) from not including the discovery GWAS in the estimation of genetic associations may be unwelcome. Additionally, differences in participant characteristics between samples may mean that genetic variants discovered in one dataset are not the most relevant predictors of the exposure in a second dataset. Another possibility for avoiding winner’s curse bias is cross-validation, whereby a large dataset is divided into discovery and estimation subsets. This can be performed efficiently by dividing the dataset into tenths and performing discovery in 90% of the data, and then obtaining association estimates in the remaining tenth (33). By repeating this procedure for each tenth of the data separately and combining results, participant overlap can be avoided while minimizing loss of power. A further suggestion is to perform a sensitivity analysis only including variants that are statistically strongly associated with the exposure (say, those that achieve *p* < 10^−11^). However, this may again lead to an unwelcome loss of power. Alternatively, a method that attempts to correct for winner’s curse bias can be employed (14–18).

Avoiding participant overlap between the discovery GWAS and the estimation dataset for associations with the outcome is an important factor in determining how to perform a Mendelian randomization analysis (28). However, pragmatic choices often have to be made to balance the possibility of a somewhat biased analysis versus the possibility of an uninformative analysis due to low sample size. Additionally, there are several other biases that could affect a Mendelian randomization analysis, the most important of which is pleiotropy (or more generally, violation of instrument validity). While analysts and reviewers should pay attention to all potential sources of bias, pleiotropy is the most critical consideration when assessing the validity of a Mendelian randomization investigation (28). Our investigation suggests that winner’s curse is a relevant consideration when deciding how to choose datasets for analysis, but winner’s curse may not bias estimates substantially or affect overall conclusions.

In conclusion, bias due to winner’s curse affects Mendelian randomization estimates, and the magnitude of bias in our empirical investigation was similar in magnitude, but generally larger than that from weak instruments. Analysts should carefully consider the possibility of avoiding sample overlap between the discovery GWAS and the estimation dataset for associations with the outcome, particularly if most genetic variants are close to the statistical significance threshold, as well as performing sensitivity analyses that reduce winner’s curse bias.

## Data Availability

UK Biobank data is available by application at https://www.ukbiobank.ac.uk/enable-your-research/apply-for-access to any bona fide researcher.

## Ethics approval

This research was conducted according to the principles expressed in the Declaration of Helsinki. The UK Biobank cohort has been approved by the North West Multicentre Research Ethics Committee, UK (Ref: 16/NW/0274). Written informed consent has been obtained from all study participants. The current study was approved by the UK Biobank Access Management Board under application 7439. Participants who had withdrawn consent by the time of the analysis were excluded from the dataset.

## Author contributions

Study conception and design: DG, SB; Analysis: TJ; writing the manuscript first draft: TJ; drafting the manuscript: all authors; reviewing and approving the manuscript: all authors.

## Data Availability Statement

UK Biobank data is available by application at https://www.ukbiobank.ac.uk/enable-your-research/apply-for-access to any *bona fide* researcher.

## Supplementary data

Supplementary Figure S1 is included as part of this submission.

## SUPPLEMENTARY MATERIAL

**Supplementary Figure S1:**
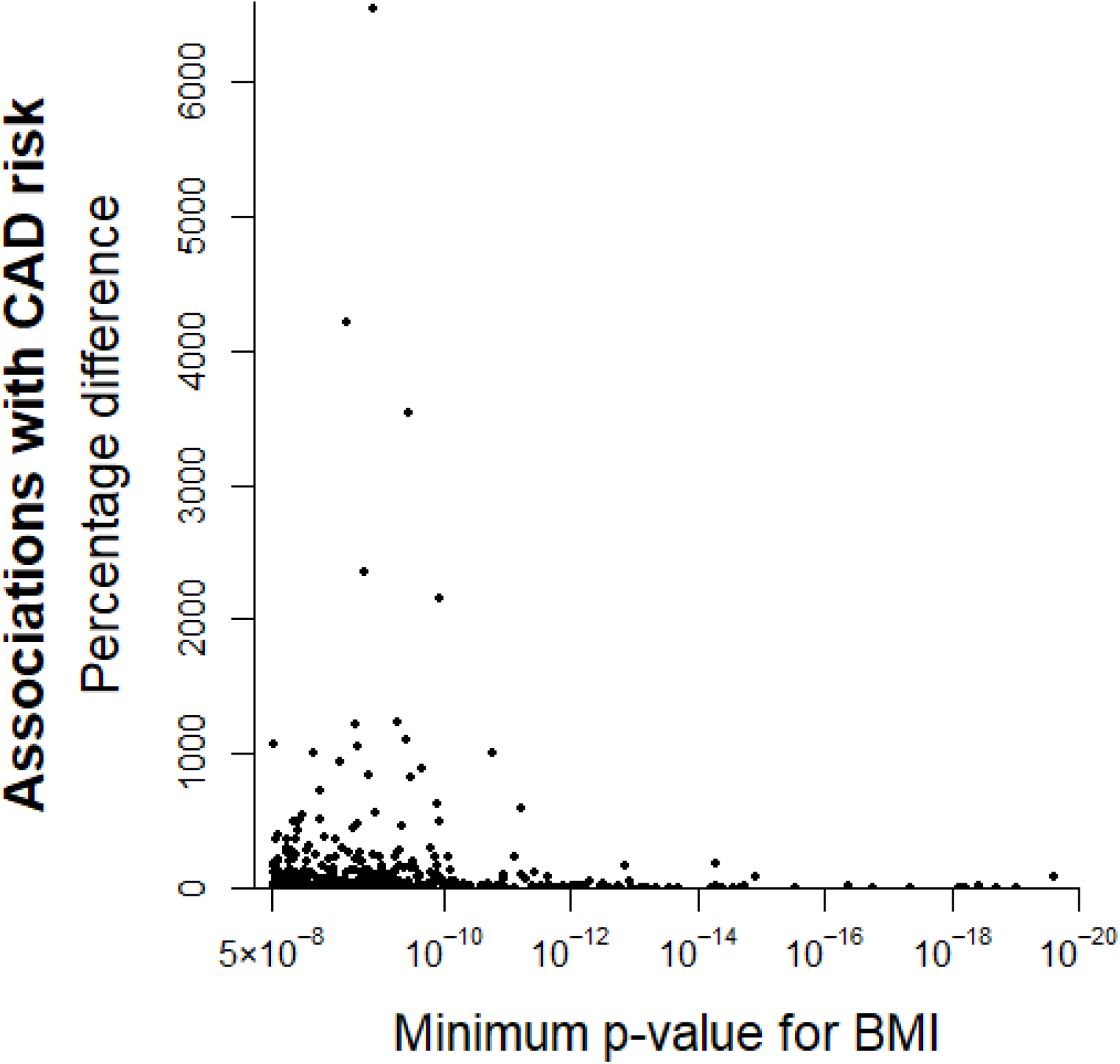
Scatter plot showing variants selected as associated with body mass index (BMI) in at least one iteration: the percentage difference in the beta-coefficient estimate for the association with coronary artery disease risk in Group A between the average value across all iterations and the average across only those iterations for which it was significant for BMI, plotted against its minimum p-value for BMI across iterations. Only one variant per locus is plotted. This plot is identical to the bottom-left panel of **Figure 1**, except the y-axis is extended here to accommodate plotting of all points.

## Bibliography

1. Lawlor DA, Harbord RM, Sterne JA, Timpson N, Davey Smith G. Mendelian randomization: using genes as instruments for making causal inferences in epidemiology. Statistics in Medicine. 2008;27(8):1133–63.

2. Burgess S, Thompson SG. Mendelian randomization: methods for using genetic variants in causal estimation: Chapman & Hall/CRC Press; 2015.

3. Hingorani A, Humphries S. Nature’s randomised trials. The Lancet. 2005;366(9501):1906–8.

4. Thanassoulis G, O’Donnell CJ. Mendelian randomization: nature’s randomized trial in the post-genome era. JAMA. 2009;301(22):2386–8.

5. Davey Smith G, Lawlor D, Harbord RM, Timpson N, Day I, Ebrahim S. Clustered environments and randomized genes: a fundamental distinction between conventional and genetic epidemiology. PLoS Medicine. 2007;4(12):e352.

6. Taylor M, Tansey KE, Lawlor DA, Bowden J, Evans DM, Davey Smith G, et al. Testing the principles of Mendelian randomization: Opportunities and complications on a genomewide scale. bioRxiv. 2017;124362.

7. Swanson SA, Tiemeier H, Ikram MA, Hernán MA. Nature as a trialist?: Deconstructing the analogy between Mendelian randomization and randomized trials. Epidemiology. 2017;28(5):653–9.

8. Visscher PM, Wray NR, Zhang Q, Sklar P, McCarthy MI, Brown MA, et al. 10 years of GWAS discovery: biology, function, and translation. American Journal of Human Genetics. 2017;101(1):5–22.

9. Burgess S, Scott RA, Timpson NJ, Davey Smith G, Thompson SG, EPIC-InterAct Consortium. Using published data in Mendelian randomization: a blueprint for efficient identification of causal risk factors. European Journal of Epidemiology. 2015;30(7):543–52.

10. Bazerman MH, Samuelson WF. I won the auction but don’t want the prize. Journal of Conflict Resolution. 1983;27(4):618–34.

11. Lohmueller KE, Pearce CL, Pike M, Lander ES, Hirschhorn JN. Meta-analysis of genetic association studies supports a contribution of common variants to susceptibility to common disease. Nature Genetics. 2003;33(2):177–82.

12. Didelez V, Sheehan N. Mendelian randomization as an instrumental variable approach to causal inference. Statistical Methods in Medical Research. 2007;16(4):309–30.

13. Burgess S, Butterworth A, Thompson SG. Mendelian randomization analysis with multiple genetic variants using summarized data. Genetic Epidemiology. 2013;37(7):658–65.

14. Bigdeli TB, Lee D, Webb BT, Riley BP, Vladimirov VI, Fanous AH, et al. A simple yet accurate correction for winner’s curse can predict signals discovered in much larger genome scans. Bioinformatics. 2016;32(17):2598–603.

15. Xiao R, Boehnke M. Quantifying and correcting for the winner’s curse in genetic association studies. Genetic Epidemiology. 2009;33(5):453–62.

16. Zhong H, Prentice RL. Correcting “winner’s curse” in odds ratios from genomewide association findings for major complex human diseases. Genetic Epidemiology. 2010;34(1):78–91.

17. Mounier N, Kutalik Z. Bias correction for inverse variance weighting Mendelian randomization. bioRxiv. 2021:2021.03.26.437168.

18. Bowden J, Dudbridge F. Unbiased estimation of odds ratios: combining genomewide association scans with replication studies. Genetic Epidemiology. 2009;33(5):406–18.

19. Nelson CR, Startz R. The distribution of the instrumental variables estimator and its t-ratio when the instrument is a poor one. Journal of Business. 1990;63(1):125–40.

20. Burgess S, Davies NM, Thompson SG. Bias due to participant overlap in two-sample Mendelian randomization. Genetic Epidemiology. 2016;40(7):597–608.

21. Pierce BL, Burgess S. Efficient design for Mendelian randomization studies: subsample and 2-sample instrumental variable estimators. American Journal of Epidemiology. 2013;178(7):1177–84.

22. Stock JH, Wright JH, Yogo M. A survey of weak instruments and weak identification in generalized method of moments. Journal of Business and Economic Statistics. 2002;20(4):518–29.

23. Burgess S, Thompson SG. Bias in causal estimates from Mendelian randomization studies with weak instruments. Statistics in Medicine. 2011;30(11):1312–23.

24. Hägg S, Fall T, Ploner A, Mägi R, Fischer K, Draisma HHM, et al. Adiposity as a cause of cardiovascular disease: a Mendelian randomization study. International Journal of Epidemiology. 2015;44(2):578–86.

25. Larsson SC, Bäck M, Rees JM, Mason AM, Burgess S. Body mass index and body composition in relation to 14 cardiovascular conditions in UK Biobank: a Mendelian randomization study. European Heart Journal. 2020;41(2):221–6.

26. Astle WJ, Elding H, Jiang T, Allen D, Ruklisa D, Mann AL, et al. The allelic landscape of human blood cell trait variation and links to common complex disease. Cell. 2016;167(5):1415–29.

27. Howie BN, Donnelly P, Marchini J. A flexible and accurate genotype imputation method for the next generation of genome-wide association studies. PLoS Genetics. 2009;5(6):e1000529.

28. Burgess S, Davey Smith G, Davies NM, Dudbridge F, Gill D, Glymour MM, et al. Guidelines for performing Mendelian randomization investigations. Wellcome Open Research. 2020;4:186.

29. Marchini J, Howie B, Myers S, McVean G, Donnelly P. A new multipoint method for genome-wide association studies by imputation of genotypes. Nature Genetics. 2007;39(7):906–13.

30. Yavorska OO, Burgess S. MendelianRandomization: an R package for performing Mendelian randomization analyses using summarized data. International Journal of Epidemiology. 2017;46(6):1734–9.

31. R Core Team. R: A language and environment for statistical computing. Vienna, Austria: R Foundation for Statistical Computing; 2021.

32. Zhao Q, Chen Y, Wang J, Small DS. Powerful three-sample genome-wide design and robust statistical inference in summary-data Mendelian randomization. International Journal of Epidemiology. 2019;48(5):1478–92.

33. Denault WR, Bohlin J, Page CM, Burgess S, Jugessur A. Cross-fitted instrument: a blueprint for one-sample Mendelian Randomization. PLOS Computational Biology. 2022.

